# Therapeutic Potential of Transcranial Magnetic Stimulation (TMS) in Long COVID: Protocol for a Systematic Review and Meta-Analysis

**DOI:** 10.1101/2025.06.05.25329096

**Authors:** Nilihan E.M. Sanal-Hayes, Kate Slade, Marie Mclaughlin, Ethan Berry, Emma Swift, Lawrence D. Hayes

## Abstract

**Objective:** To synthesise quantitative evidence on the effectiveness of Transcranial Magnetic Stimulation (TMS) as an intervention for individuals with long COVID/post-COVID syndrome.

**Introduction:** An estimated 65 million people worldwide meet the WHO’s criteria for post-COVID-19 condition, a multisystem disorder with persistent symptoms following SARS-CoV-2 infection. Despite its global impact, effective treatments are limited. A recent review highlighted early but promising results from non-invasive brain stimulation techniques, including transcranial magnetic stimulation (TMS), though the underlying mechanisms remain unclear. Given the expanding use of TMS in long COVID, a follow-up meta-analysis is needed to reflect recent developments. This review aims to evaluate the efficacy of TMS in treating post-COVID-19 symptoms.

**Inclusion criteria:** This review will include studies that investigate interventions involving TMSin individuals diagnosed with long COVID or post-COVID syndrome, limited to those published in the English language. Studies will be excluded if they do not involve rTMS as a therapeutic intervention, do not involve TMS at all, focus on populations other than those with long COVID or post-COVID condition, are review articles or case studies, or are not published in English.

**Methods:** A comprehensive search will be conducted in CINAHL Ultimate, MEDLINE, ScienceDirect, and Scopus using a strategy developed with the research team. Retrieved citations will be managed in Rayyan. Two independent reviewers will screen titles and abstracts, followed by full-text screening and data extraction by two other independent reviewers. All stages will follow predefined, pilot-tested inclusion and exclusion criteria. Discrepancies will be resolved by a third reviewer or team discussion. Intra-rater reliability at both screening stages will be assessed using Cohen’s Kappa. Study characteristics and findings will be presented using both narrative synthesis and tabular formats.

**Review registration:** https://doi.org/10.17605/OSF.IO/RE235

## Introduction

According to the NICE guidelines, individuals experiencing post-viral symptoms lasting from 4 weeks to over 12 weeks following acute COVID-19 infection are considered to have a condition known as long COVID (4). An estimated 65 million individuals worldwide meet the diagnostic criteria for post-COVID-19 condition, as defined by the World Health Organization (WHO), a multisystem disorder characterised by a diverse range of persistent symptoms following acute infection with SARS-CoV-2 (1). This condition is characterised by a wide range of symptoms, over 100 have been identified including muscle pain, severe fatigue, and cognitive impairments, with significant symptom overlap observed with Myalgic Encephalomyelitis/Chronic Fatigue Syndrome (ME/CFS) (5).

Previous studies have demonstrated that individuals with long COVID experience a range of debilitating symptoms, including pain, fatigue, post-exertional malaise, sleep disturbances, breathlessness, and neurological abnormalities (5–12). Among these, musculoskeletal pain has been identified as the most prevalent persistent symptom in individuals with long COVID (6). Fatigue is another hallmark symptom. Approximately 46% of individuals with long COVID report persistent fatigue lasting from weeks to months (7). Post-exertional malaise, a prolonged worsening of symptoms following minimal physical or mental activity is also a key feature in this condition. One study found that 59% of individuals with long COVID met the criteria for post-exertional malaise used in ME/CFS research (8). Sleep disturbances are also highly prevalent. In long COVID, approximately 65% of patients report poor sleep quality (9). Breathlessness is another frequently reported symptomand has been identified as particularly debilitating (10). Kim et al. (11)reported a potential association between persistent breathlessness and small airway functional impairment in individuals with long COVID. Neurological abnormalities are documented, and Pilotto et al. (12) reported that 40% of hospitalised and non-hospitalised COVID-19 patients exhibited neurological abnormalities during examination such as hyposmia, cognitive deficits, postural tremor and subtle motor/sensory deficits.

Other frequently reported well-being outcomes associated with long COVID include reduced quality of life, as well as symptoms of anxiety and depression (13–15). Health-related quality of life has been shown to be significantly lower in individuals with a history of COVID-19 infection (13). Symptoms of anxiety and depression are also prevalent and have been observed as early as three months post-infection (14). However, findings remain inconsistent, with some studies reporting elevated psychological distress, while others indicate variability depending on symptom severity and duration (14). Despite the global burden and complex symptomatology of long COVID, effective treatment options remain scarce. A recent systematic review by Markser et al. (2) synthesised current evidence on the efficacy of non-invasive and minimally invasive brain stimulation techniques including TMS for alleviating symptoms associated with post-COVID-19 syndrome.

Transcranial magnetic stimulation (TMS) is a non-invasive brain stimulation technique that uses a coil placed on the scalp to deliver magnetic pulses. Through electromagnetic induction, these pulses generate a magnetic field that induces an electrical current in the cortical tissue beneath the coil (16). The effects of TMS can be either acute or prolonged, depending on stimulation parameters such as intensity, coil shape and orientation, and the frequency and pattern of pulses. Single-pulse TMS is primarily used to investigate brain function. For instance, delivering a single pulse over the primary motor cortex (M1) can evoke motor responses—known as motor evoked potentials (MEPs)—in target muscles, which are measured using electromyography (EMG) (17).The amplitude and latency of these MEPs provide insights into motor cortex excitability (18). In contrast, repetitive TMS (rTMS) can modulate neuronal activity in a way that produces effects lasting beyond the stimulation period (19). The effects of rTMS on neural activity depend on the frequency and pattern of stimulation, with certain protocols producing inhibitory effects and others excitatory. Repeated sessions have been explored as treatments for various psychiatric and neurological disorders, owing to their potential to induce long-lasting changes in neural plasticity (20,21).

Markser et al. (2) reported that while existing studies show promising initial results with improvements in clinical outcome measures, the mechanistic understanding of post-COVID-19 and the potential benefits of brain stimulation techniques remains limited. Given the growing application of this technique in individuals with long COVID (3) and related conditions(22–24), a follow-up meta-analysis is warranted to reflect recent advancements and the expanding evidence base. This review aims to assess the therapeutic effectiveness of rTMS in the treatment of post-COVID-19 symptoms.

### Review questions

#### This review poses primary and secondary research questions

Primary research question: What is the effectiveness of rTMS as an intervention for individuals with long COVID/Post-COVID symptoms (e.g., fatigue, cognitive dysfunction, mood disturbances, quality of life)?

#### Secondary research questions

1. Are there differences in the effectiveness of rTMS based on stimulation parameters (e.g., frequency, intensity, targeted brain region)?
2. Do participant characteristics (e.g., age, sex, duration since acute COVID-19 infection) moderate the effectiveness of rTMS?
3. What is the risk of bias and methodological quality of studies investigating rTMS for long COVID/Post-COVID?

We approach this review with the hypothesis that rTMS may offer therapeutic benefits for individuals experiencing persistent symptoms of long COVID/Post-COVID, particularly in domains such as fatigue, depression, and cognitive dysfunction, which are commonly targeted in other neuropsychiatric conditions treated with rTMS. We also expect that, the evidence base will be limited but emerging, given the recency of long COVID as a clinical entity and the time required to conduct and publish controlled trials, and most studies will be small-scale or pilot RCTs, possibly with methodological variability in study design, outcome measures, and stimulation parameters. Moreover, we expect that symptom improvements may vary by domain, with stronger effects hypothesised for mood-related symptoms (e.g., depression) than for more complex or diffuse symptoms like fatigue or brain fog. Given the novelty and evolving definition of long COVID, we anticipate some heterogeneity in how populations are defined across studies, which may influence both inclusion decisions and the strength of conclusions.

### Eligibility Criteria

#### Participants

This review will investigate interventions involving rTMS in individuals diagnosed with long COVID or post-COVID syndrome living, without imposing any demographic restrictions.

#### Concept

The concepts in the review include rTMS and long COVID.

*Transcranial Magnetic Stimulation* is a non-invasive brain stimulation technique that uses a coil placed on the scalp to deliver magnetic pulses. Through electromagnetic induction, these pulses generate a magnetic field that induces an electrical current in the cortical tissue beneath the coil (16).

*Long COVID* is a condition characterised by post-viral symptoms that persist for at least 4 weeks and can last beyond 12 weeks following an acute COVID-19 infection (4). This condition is characterised by a wide range of symptoms, over 100 have been identified including muscle pain, severe fatigue, and cognitive impairments, with significant symptom overlap observed with Myalgic Encephalomyelitis/Chronic Fatigue Syndrome (ME/CFS) (5).

We will exclude articles that use TMS for purposes other than therapeutic intervention, (b) do not involve rTMS, (c) focus on populations other than those with long COVID/post-COVID condition, (d) are review articles or case studies, or (e) are not published in English. We will include grey literature alongside peer-reviewed sources. This includes preprints (e.g., PsyArXiv, arXiv), dissertations (via ProQuest), conference proceedings (e.g., Web of Science), and institutional repositories such as DSpace. Furthermore, we will supplement database searches by reviewing reference lists (forward citation searching) and identifying articles that cite included studies (backward citation searching) using tools such as CrossRef and Google Scholar. We will also use CoCites, if available, to find articles with similar citation patterns.

### Context

Articles must include individuals with long COVID or be described using relevant terms that indicate this condition. Repetitive Transcranial Magnetic Stimulation must be used specifically as a treatment or intervention. Articles must present at least pilot data; therefore, review articles and case studies should not be included. Articles must be written in English and include quantitative data.

### Types of Sources

This review will consider articles that present at least pilot data and include quantitative data. Review articles, case studies or qualitative studies will therefore not be considered.

## Methods

This meta-analysis will be conducted in accordance with the PRISMA (Preferred Reporting Items for Systematic Reviews and Meta-Analyses) guidelines. The protocol has been registered with Open Science Framework: https://doi.org/10.17605/OSF.IO/RE235. Any deviation from the protocol will be clearly logged with dates, reasons, and impacts noted.

### Search strategy

Our search strategy was designed to achieve a balance between comprehensive coverage of the relevant literature and practical constraints, while maintaining a high level of scientific rigour. We selected four databases—CINAHL Ultimate, MEDLINE, ScienceDirect, and Scopus—based on their relevance to the interdisciplinary nature of our topic, which spans psychology, health sciences, and behavioural research. CINAHL Ultimate was chosen for its strong coverage of nursing and allied health literature. MEDLINE (accessed via EBSCOhost) provides authoritative biomedical literature, including psychiatry and clinical psychology studies. ScienceDirect was included due to its extensive full-text access to journals in psychology, neuroscience, and related disciplines published by Elsevier. Scopus was selected for its comprehensive indexing of interdisciplinary research and citation tracking capabilities, allowing us to identify additional relevant studies. We will search grey literature alongside peer-reviewed sources. This includes preprints (e.g., PsyArXiv, arXiv), dissertations (via ProQuest), conference proceedings (e.g., Web of Science), and institutional repositories like DSpace. Moreover, we will supplement database searches by reviewing reference lists, identifying citing articles via tools such as CrossRef and Google Scholar, and using CoCites to find articles with similar citation patterns. We will validate our search by testing it against known relevant studies to ensure key articles are captured. If too many irrelevant results appear, we will refine the search terms or criteria. This iterative process will continue until the strategy is optimised for relevance and accuracy. We will contact study authors via a standardised email when additional details are needed for eligibility or analysis. Follow-up emails will be sent after two and an additional two weeks if there’s no response.

All contact attempts and responses will be documented. To ensure transparency, we will report how many authors were contacted, how many responded, and how many provided the requested data. These details will appear in the main manuscript and in a supplementary table summarising the information requested and obtained. We do not plan a living review but will repeat database searches before submission if over six months have passed since the original search.

### Query strings

TI ((long COVID OR post-COVID OR “post-acute sequelae of SARS-CoV-2 infection” OR “post-viral fatigue” OR “COVID-19 recovery syndrome” OR “long-haul COVID” OR “long COVID syndrome”)) OR AB ((long COVID OR post-COVID OR “post-acute sequelae of SARS-CoV-2 infection” OR “post-viral fatigue” OR “COVID-19 recovery syndrome” OR “long-haul COVID” OR “long COVID syndrome”))

This review focuses on clinical and functional outcomes related to long COVID/Post-COVID symptoms following Repetitive Transcranial Magnetic Stimulation (rTMS). Key outcomes include fatigue, cognitive function, mood/psychological symptoms, sleep disturbances, quality of life, and overall symptom improvement. We will also gather data on adverse effects, TMS tolerability, and treatment durability. The independent variable is rTMS as a treatment for long COVID/Post-COVID symptoms, particularly fatigue, cognitive dysfunction, depression, and anxiety. Studies with various TMS protocols will be included, such as high-frequency TMS (≥10 Hz), low-frequency TMS (1 Hz), theta burst stimulation (TBS), and repetitive TMS (rTMS), with different stimulation targets (e.g., dorsolateral prefrontal cortex) and treatment durations. Comparator groups may include sham TMS, standard care, or no-treatment controls. We will also analyse variables that may influence treatment outcomes, including participant characteristics (age, sex, time since infection, COVID-19 severity, comorbid conditions), study-level factors (sample size, design, risk of bias, study setting), TMS protocol variables (site, frequency, intensity, session number), and follow-up duration.

### Study/Source of Evidence Selection

After the search, all citations will be uploaded and have their duplicates removed in Rayyan (25). The screening process will consist of two rounds: (1) title and abstract screening, and (2) full-text screening. In both rounds, two independent reviewers will screen each record using pre-specified inclusion and exclusion criteria developed a priori. Blinding of reviewers during screening will be implemented to the extent possible using Rayyan, which allows for independent decisions without visibility into the other reviewer’s judgments. This helps reduce bias and increases objectivity in the initial phases of study selection. Conflicts between reviewers will be flagged automatically by Rayyan and subsequently resolved through discussion with two other reviewers. If consensus cannot be reached, a third reviewer (independent chair) will adjudicate. To assure consistency and transparency, all reviewers will undergo a calibration exercise using a small sample of studies before formal screening begins. At both the titles and abstracts stage and the full text stage, inter-rater reliability will be calculated and expressed via Cohen’s Kappa statistic, scores range from –1 to 1 with scores closer to 1 indicating stronger agreement. This exercise will help refine the application of the inclusion/exclusion criteria and ensure a shared understanding of borderline cases. We will share the full list of sources from the database searches and screening decisions by individual screeners. Bibliographic data (titles, abstracts, metadata) will be exported in RIS and CSV formats, while screening decisions will be provided in a separate CSV/XLSX file. All files will be uploaded to an open-access repository, like OSF, upon manuscript submission or acceptance.

### Data Extraction

In the training and calibration stage, a small sample of studies (5-10%) will be independently extracted by all reviewers using the draft form, with discrepancies discussed and resolved to refine the protocol. In the primary data extraction stage, two reviewers will independently extract key data (e.g., means, standard deviations [SDs], TMS parameters, demographics, outcomes) using a standardised form. In the risk of bias (RoB) and methodological quality extraction stage, two reviewers will independently assess study-level risk of bias using tools like the Cochrane RoB tool, with discrepancies resolved through discussion. In the reconciliation and verification stage, discrepancies will be resolved through discussion or adjudication by a third reviewer, with final data entered into the meta-analysis dataset.

Optional AI/computer-assisted support may assist in identifying relevant text or extracting bibliographic metadata, but all outputs will be verified by human reviewers. All stages will be conducted by humans, with AI support supervised by humans.

Data extraction will follow PRISMA guidelines and Cochrane Handbook procedures, using the standardized form provided in the OSF project (Data_Extraction_Form_TMS_LongCOVID.xlsx). Only data related to long COVID/Post-COVID, as defined by the study authors, will be extracted, with missing data marked as “NR” and flagged for follow-up. Primary study information, including bibliographic details (author(s), year, title, journal, country, and institution), study design (e.g., RCT, quasi-experimental), and setting (e.g., clinical, academic), will be recorded. Participant characteristics such as mean age, age range, sex/gender distribution, time since acute COVID-19, severity of initial infection, and comorbidities will be extracted. Intervention details, including TMS protocol type, stimulation site, frequency, intensity, session number, neuronavigation use, and treatment duration/schedule, will also be recorded. Comparator groups, including control conditions (e.g., sham TMS, standard care, waitlist/no treatment), will be described. Clinical outcomes (e.g., fatigue, cognitive function, mood, sleep, quality of life, symptom improvement) and statistical data (e.g., means, SDs, sample sizes, p-values, effect sizes) will be extracted, along with follow-up data and adverse effects. Risk of bias will be assessed using the Cochrane Risk of Bias 2.0 tool, covering domains such as randomisation, blinding, attrition, and selective reporting. Ambiguous data will be flagged for discussion, and assumptions or clarifications will be noted in comments, with no estimation of missing data unless explicitly instructed. All numerical entries will be double-checked prior to submission, and a second reviewer will verify the data, resolving discrepancies through discussion or adjudication by a third reviewer. Completed forms will be saved and uploaded to the OSF folder following the naming convention: StudyID_ExtractorInitials_Extraction.xlsx.

Each round of data extraction will be conducted by two independent extractors working in parallel, covering study characteristics, TMS intervention details, participant data, clinical outcomes, and risk of bias assessments. Extractors will receive standardised training and work independently, with results compared and discrepancies reconciled through discussion. A third reviewer will adjudicate if disagreements persist. Inter-rater reliability will be assessed during training using Cohen’s kappa for categorical variables and ICCs for continuous variables, based on a subset of studies. These results will inform adjustments to the extraction form and training. In each data extraction round, two independent extractors will compare their entries side by side, with differences highlighted automatically. They will then discuss discrepancies, referring to source material and instructions, to reach consensus. If they cannot agree, a third senior reviewer will adjudicate, reviewing the relevant documents and providing a final decision, recorded in the reconciliation log. All discrepancies and resolutions will be documented. Systematic discrepancies will be analysed to update the extraction protocol if needed. Once reconciled, the data will be finalised for analysis and included in the meta-analytic dataset.

### Data Analysis and Presentation

The meta-analysis will focus on estimating the overall effect of rTMS on long COVID/Post-COVID symptoms, such as fatigue, cognitive dysfunction, and mood disturbances. Effect sizes from continuous outcomes will be standardised (e.g., Hedges’ g or standardised mean difference), and odds ratios will be used for dichotomous outcomes when applicable. A random-effects model will be used throughout, with analyses conducted in R Studio using the *metafor* package. Heterogeneity will be assessed via *I2, Cochran’s Q*, and *tau*^*2*^ statistics.

Subgroup analyses will explore variations in TMS protocols, participant characteristics (e.g., age, sex, COVID-19 severity), study features, and symptom domains. Moderator analyses will assess how TMS parameters and participant traits influence outcomes, and the impact of risk of bias will be tested through sensitivity analyses. Additional sensitivity checks will examine the effects of excluding high-risk studies, handling missing data, and methodological quality. Where available, follow-up data will be used to assess treatment durability, and adverse effects will be summarised qualitatively or quantitatively. If data allow, meta-regression will be used to examine relationships between continuous variables (e.g., number of sessions) and effect sizes. In cases of missing or insufficient data, planned analyses will be adjusted and limitations clearly reported. Results will be presented using forest and funnel plots, and a GRADE profile will summarise the quality of evidence for each outcome. Conclusions will be based on α level, confidence intervals, and effect sizes. Effect sizes will be interpreted using thresholds: Cohen’s d ≥ 0.20 as small but meaningful, ≥ 0.50 as moderate, and ≥ 0.80 as large; ORs or RRs ≥ 1.5 will indicate clinical relevance for dichotomous outcomes. High heterogeneity (*I^2^* > 50%) will prompt cautious interpretation and exploration through subgroup or moderator analyses. The GRADE framework will guide the confidence in conclusions, depending on evidence quality. For rTMS to be considered clinically useful for long COVID, we expect consistent small-to-moderate effects across key outcomes like fatigue and cognitive dysfunction. If subgroup findings show stronger effects in specific populations or symptom domains, these will be highlighted. Adverse effects, if present in ≥10% of participants, will temper conclusions. If a saturation point is reached (5– 10 studies) and new data do not significantly shift results, findings will be considered stable. Long-term follow-up data (≥3 months) will strengthen conclusions on durability. Lastly, evidence of publication bias will lead to more cautious interpretation regarding the generalisability of results.

## Discussion

This meta-analysis protocol aims to assess the therapeutic effectiveness of rTMS in the treatment of post-COVID-19 symptoms. With growing application of these techniques, particularly rTMS, in individuals with long COVID (3) and related conditions (22–24), an up-to-date meta-analysis is warranted. The decision to conduct a meta-analysis arose from the need to identify key symptom outcomes of rTMS, including parameters such as frequency, intensity, safety, target sites, and existing gaps in the current literature. This protocol is not a critical appraisal of studies due to the emerging nature of the field but aims to provide an overview of the available research. The review will highlight the efficacy of TMS in managing long COVID symptoms like fatigue, cognitive dysfunction, and mood disturbances, helping to inform future research directions and clinical practice. By providing context to current findings, this research can guide future studies to enhance treatment effectiveness for long COVID.

### Project Timeline

The estimated project timeline includes screening of titles, abstracts, and full texts from June to August 2025, followed by data extraction from August to October 2025. Results are expected by November 2025. During this period, we will update our database searches to identify any newly published studies that meet our inclusion criteria.

## Acknowledgements

None.

## Data availability

This is a protocol paper, so no data are currently available. Once the study is complete, the dataset supporting the findings will be included in the main manuscript and Appendix.

## Authors contributions

All authors contributed equally to the conceptualisation, preliminary search, search strategy, writing – original draft, review and editing

## References

1. Davis HE, McCorkell L, Vogel JM, Topol EJ. Long COVID: major findings, mechanisms and recommendations. Nat Rev Microbiol. 2023;21(3):133–46.

2. Markser A, Vöckel J, Schneider A, Baumeister-Lingens L, Sigrist C, Koenig J. Non-Invasive Brain Stimulation for Post-COVID-19 Conditions: A Systematic Review. Am J Med. 2025 Apr;138(4):681–97.

3. Sasaki N, Yamatoku M, Tsuchida T, Sato H, Yamaguchi K. Effect of Repetitive Transcranial Magnetic Stimulation on Long Coronavirus Disease 2019 with Fatigue and Cognitive Dysfunction. Prog Rehabil Med. 2023;8:20230004.

4. Raveendran AV, Jayadevan R, Sashidharan S. Long COVID: An overview. Diabetes Metab Syndr. 2021;15(3):869–75.

5. Hayes LD, Ingram J, Sculthorpe NF. More Than 100 Persistent Symptoms of SARS-CoV-2 (Long COVID): A Scoping Review. Frontiers in Medicine [Internet]. 2021 [cited 2024 Feb 15];8. Available from: https://www.frontiersin.org/articles/10.3389/fmed.2021.750378

6. Khoja O, Silva Passadouro B, Mulvey M, Delis I, Astill S, Tan AL, et al. Clinical Characteristics and Mechanisms of Musculoskeletal Pain in Long COVID. Journal of Pain Research. 2022 Jun 17;15:1729–48.

7. Sandler CX, Wyller VBB, Moss-Morris R, Buchwald D, Crawley E, Hautvast J, et al. Long COVID and Post-infective Fatigue Syndrome: A Review. Open Forum Infect Dis. 2021 Oct;8(10):ofab440.

8. Twomey R, DeMars J, Franklin K, Culos-Reed SN, Weatherald J, Wrightson JG. Chronic Fatigue and Postexertional Malaise in People Living With Long COVID: An Observational Study. Phys Ther. 2022 Apr 1;102(4).

9. Gui Z, Wang YY, Li JX, Li XH, Su Z, Cheung T, et al. Prevalence of poor sleep quality in COVID-19 patients: a systematic review and meta-analysis. Front Psychiatry. 2024 Jan 8;14:1272812.

10. Zheng B, Daines L, Han Q, Hurst JR, Pfeffer P, Shankar-Hari M, et al. Prevalence, risk factors and treatments for post-COVID-19 breathlessness: a systematic review and meta-analysis. European Respiratory Review [Internet]. 2022 Dec 31 [cited 2024 Feb 8];31(166). Available from: https://err.ersjournals.com/content/31/166/220071

11. Kim M, Hwang J, Grist JT, Abueid G, Yoon SH, Grau V, et al. Functional Impairment in Small Airways Associated With the Breathlessness Symptoms in Long–Coronavirus Disease. Journal of Thoracic Imaging. :10.1097/RTI.0000000000000748.

12. Pilotto A, Cristillo V, Cotti Piccinelli S, Zoppi N, Bonzi G, Sattin D, et al. Long-term neurological manifestations of COVID-19: prevalence and predictive factors. Neurol Sci. 2021 Dec 1;42(12):4903–7.

13. Arab-Zozani M, Hashemi F, Safari H, Yousefi M, Ameri H. Health-Related Quality of Life and its Associated Factors in COVID-19 Patients. Osong Public Health Res Perspect. 2020 Oct;11(5):296–302.

14. Shanbehzadeh S, Tavahomi M, Zanjari N, Ebrahimi-Takamjani I, Amiri-arimi S. Physical and mental health complications post-COVID-19: Scoping review. Journal of Psychosomatic Research. 2021 Aug 1;147:110525.

15. Means-Christensen AJ, Roy-Byrne PP, Sherbourne CD, Craske MG, Stein MB. Relationships among pain, anxiety, and depression in primary care. Depression and Anxiety. 2008;25(7):593–600.

16. Rossini PM, Burke D, Chen R, Cohen LG, Daskalakis Z, Di Iorio R, et al. Non-invasive electrical and magnetic stimulation of the brain, spinal cord, roots and peripheral nerves: Basic principles and procedures for routine clinical and research application. An updated report from an I.F.C.N. Committee. Clinical Neurophysiology. 2015 Jun 1;126(6):1071–107.

17. Hallett M. Transcranial Magnetic Stimulation: A Primer. Neuron. 2007 Jul 19;55(2):187–99.

18. Nuttall HE, Kennedy-Higgins D, Hogan J, Devlin JT, Adank P. The effect of speech distortion on the excitability of articulatory motor cortex. NeuroImage. 2016 Mar 1;128:218–26.

19. Ziemann U. TMS in cognitive neuroscience: Virtual lesion and beyond. Cortex. 2010 Jan 1;46(1):124–7.

20. O’Reardon JP, Solvason HB, Janicak PG, Sampson S, Isenberg KE, Nahas Z, et al. Efficacy and safety of transcranial magnetic stimulation in the acute treatment of major depression: a multisite randomized controlled trial. Biol Psychiatry. 2007 Dec 1;62(11):1208–16.

21. Morriss R, Briley PM, Webster L, Abdelghani M, Barber S, Bates P, et al. Connectivity-guided intermittent theta burst versus repetitive transcranial magnetic stimulation for treatment-resistant depression: a randomized controlled trial. Nat Med. 2024 Feb;30(2):403–13.

22. Miwa K, Inoue Y. Repetitive transcranial magnetic stimulation ameliorates symptoms in patients with myalgic encephalomyelitis (chronic fatigue syndrome). IBRO Neuroscience Reports. 2023;15:335–41.

23. Yang DG, Gu R, Kubo J, Kakuda W. Is the efficacy of repetitive transcranial magnetic stimulation influenced by baseline severity of fatigue symptom in patients with myalgic encephalomyelitis. The International journal of neuroscience. 2020 Jan;130(1):64–70.

24. Kakuda W, Momosaki R, Yamada N, Abo M. High-frequency rTMS for the Treatment of Chronic Fatigue Syndrome: A Case Series. Internal medicine (Tokyo, Japan). 2016 Jan;55(23):3515–9.

25. Ouzzani M, Hammady H, Fedorowicz Z, Elmagarmid A. Rayyan—a web and mobile app for systematic reviews. Systematic Reviews. 2016 Dec 5;5(1):210.

